# Neurological manifestations associated with COVID-19: a nationwide registry

**DOI:** 10.1101/2020.07.15.20154260

**Authors:** Elodie Meppiel, Nathan Peiffer-Smadja, Alexandra Maury, Imen Bekri, Cécile Delorme, Virginie Desestret, Lucas Gorza, Geoffroy Hautecloque-Raysz, Sophie Landre, Annie Lannuzel, Solène Moulin, Peggy Perrin, Paul Petitgas, François Sellal, Adrien Wang, Pierre Tattevin, Thomas de Broucker, on behalf of the contributors to the NeuroCOVID registry

**Affiliations:** Department of Neurology, Centre Hospitalier de Saint-Denis, Hôpital Delafontaine, F-93200 Saint-Denis, France; Department of Infectious Diseases, Bichat-Claude Bernard Hospital, Assistance-Publique Hôpitaux de Paris, F-75018 Paris, France; Université de Paris, IAME, INSERM, F-75018 Paris, France; Department of Neurology and Stroke Center, Centre Hospitalier de Versailles, F-78150 Le Chesnay, France; Department of Neurology, Pitié-Salpêtrière Hospital, Assistance-Publique Hôpitaux de Paris, F-75013 Paris, France; Department of Neuro-cognition and Neuro-ophthalmology, Hospices Civils de Lyon, Pierre Wertheimer Neurological Hospital, Lyon, France; the University of Lyon, Claude Bernard Lyon 1 University, F-69000 Lyon, France; Department of Neurology, Hôpital Foch, F-92151 Suresnes, France; Department of Neurology, Hôpitaux civils de Colmar, 68024 COLMAR, France; Department of Infectious Diseases, Hospices Civils de Lyon; the University of Lyon, Claude Bernard Lyon 1 University, F-69000 Lyon, France; Department of Neurology, Centre Hospitalier Universitaire de la Guadeloupe, Faculté de médecine de l’université des Antilles, Centre d’investigation clinique Antilles Guyane, Inserm CIC 1424, F-97100 Pointe-à-Pitre, France; Department of Neurology, Centre Hospitalier Universitaire, Hôpital Maison Blanche, F-51092 Reims, France; Department of Nephrology, Hôpitaux Universitaires de Strasbourg, F-67091 Strasbourg, France; Department of Infectious Diseases and Intensive Care Medicine, Centre Hospitalier Universitaire de Rennes, F-35000 Rennes, France

**Keywords:** COVID-19, SARS-CoV-2, Nervous System, Neurological manifestations, Registry

## Abstract

**Background:** The clinical description of the neurological manifestations in COVID-19 patients is still underway. This study aims to provide an overview of the spectrum, characteristics and outcomes of neurological manifestations associated with SARS-CoV-2 infection.

**Methods:** We conducted a nationwide, multicentric, retrospective study during the French COVID-19 epidemic in March-April 2020. All COVID-19 patients with *de novo* neurological manifestations were eligible.

**Results:** We included 222 COVID-19 patients with neurological manifestations from 46 centers throughout the country. Median age was 65 years (IQR 53-72), and 136 patients (61.3%) were male. COVID-19 was severe or critical in almost half of the patients (102, 45.2%). The most common neurological diseases were COVID-19 associated encephalopathy (67/222, 30.2%), acute ischemic cerebrovascular syndrome (57/222, 25.7%), encephalitis (21/222, 9.5%), and Guillain-Barré Syndrome (15/222, 6.8%). Neurological manifestations appeared after first COVID-19 symptoms with a median (IQR) delay of 6 (3-8) days in COVID-19 associated encephalopathy, 7 (5-10) days in encephalitis, 12 (7-18) days in acute ischemic cerebrovascular syndrome and 18 (15-28) days in Guillain-Barré Syndrome. Brain imaging was performed in 192 patients (86.5%), including 157 MRI (70.7%). Brain MRI of encephalitis patients showed heterogeneous acute non vascular lesion in 14/21 patients (66.7%) with associated small ischemic lesion or microhemorrhages in 4 patients. Among patients with acute ischemic cerebrovascular syndrome, 13/57 (22.8%) had multi territory ischemic strokes, with large vessel thrombosis in 16/57 (28.1%). Cerebrospinal fluid was analyzed in 97 patients (43.7%), with pleocytosis in 18 patients (18.6%). A SARS-CoV-2 PCR was performed in 75 patients and was positive only in 2 encephalitis patients. Among patients with encephalitis, ten out of 21 (47.6%) fully recovered, 3 of whom received corticosteroids (CS). Less common neurological manifestations included isolated seizure (8/222, 3.6%), critical illness neuropathy (8/222, 3.6%), transient alteration of consciousness (5/222, 2.3%), intracranial hemorrhage (5/222, 2.3%), acute benign lymphocytic meningitis (3/222, 1.4%), cranial neuropathy (3/222, 1.4%), single acute demyelinating lesion (2/222, 0.9%), Tapia syndrome (2/222, 0.9%), cerebral venous thrombosis (1/222, 0.5%), sudden paraparesis (1/222, 0.5%), generalized myoclonus and cerebellar ataxia (1/222, 0.5%), bilateral fibular palsy (1/222, 0.5%) and isolated neurological symptoms (headache, anosmia, dizziness, sensitive or auditive symptoms, hiccups, 15/222, 6.8%). The median (IQR) follow-up of the 222 patients was 24 (17-34) days with a high short-term mortality rate (28/222, 12.6%).

**Conclusion:** Neurological manifestations associated with COVID-19 mainly included CAE, AICS, encephalitis and GBS. Clinical spectrum and outcomes were broad and heterogeneous, suggesting different underlying pathogenic processes.

## INTRODUCTION

COVID-19, the disease linked to the SARS-CoV-2 coronavirus, is an emerging infectious disease with first cases reported in China in December 2019. Since then, the virus has continued to spread and on March 11, 2020 the World Health Organization (WHO) characterized COVID-19 as a pandemic. France has been heavily affected by the COVID-19 epidemic and went into lockdown on March 17, 2020. From March 1, 2020 to April 30,2020, 89 818 new cases of COVID-19 were hospitalized in France. Up to July 2, 2020, more than 160,000 cases and 29,000 deaths had been reported in France. Although common manifestations of the disease including respiratory tract and associated systemic manifestations have been well described^1,2^, neurological manifestations elicited few communications. Headaches, dizziness, anosmia, encephalopathy and strokes have been reported in cohort studies ^3,4^. However, the potential pathogenesis of SARS-CoV-2 in the central nervous system remains unclear^5^, and the range of neurological disorders associated with COVID-19 is not fully defined. The present study aims to provide a comprehensive overview of neurological manifestations associated with SARS-CoV-2 infection and to describe the clinical course and outcomes of COVID-19 patients with neurological manifestations.

## METHODS

### Study design

We conducted a nationwide retrospective, multicentric, observational study to collect neurological manifestations associated with COVID-19. A case report form (CRF) containing the main information regarding COVID-19 and neurological manifestations was sent at regular intervals from March 16 to April 27, 2020 to neurologists, infectious diseases specialists and intensivists through the National College of General Hospital Neurologists (CNNHG), the French Society for Infectious Diseases (SPILF), the Association of French-Speaking Liberal Neurologists (ANLLF), and the Multiple Sclerosis Society (SF-SEP). The anonymized CRF were collected by email or telefax at the Neurology department of Saint-Denis Hospital, France. In accordance with French law, the study complied with the *Commission Nationale de l’Informatique et des Libertés* (CNIL, reference number 2217844) and ethics committee (reference number RCB 2020-A01300-39) requirements. The IRB CER-MIT n° 2020-0602 COVID has approved the study.

### Patients and data collection

We included COVID-19 adult patients with any neurological manifestations occurring 5 days before to 35 days after the first symptoms of COVID-19. COVID-19 patients managed in intensive care units with neurological manifestations after withdrawal of sedation were also included. Only *de novo* neurological symptoms were considered, excluding patients with exacerbations of chronic neurological diseases. A confirmed case of COVID-19 was defined as a patient with a positive SARS-CoV-2 RT-PCR assay on a naso-pharyngeal sample or a positive SARS-CoV-2 serology (according to local laboratory standard). As RT-PCR analysis and serology were unavailable in some centers during the epidemic, some cases were considered COVID-19 if the clinical history and the chest CT-scan were considered typical according to local expert clinicians. We defined the severity of COVID-19 illness as mild, moderate, severe and critical according to NIH guidelines ^6^. Demographical and clinical data, neurological workup findings and outcome were systematically collected. If necessary, in-charge clinicians were directly contacted to collect any missing or uncertain records.

### Classification of neurological manifestations

Neurological manifestations were split into Central Nervous System (CNS) and Peripheral Nervous System (PNS) manifestations. Each manifestation was then classified into the following diagnostic categories:

- Stroke was considered in patients with sudden neurological deficit related to an acute vascular lesion on cerebral MRI or CT scan (ischemic and hemorrhagic stroke), in patients with transient focal deficit and normal MRI (transient ischemic attack), or in patients with cerebral venous thrombosis.
- Encephalitis was defined as altered mental status lasting ≥ 24 hours and one of the following criteria: white blood count cells (WBCs) in cerebrospinal fluid (CSF) > 5/mm^3^; presence of compatible acute lesion on brain MRI.
- Encephalopathy was defined by delirium lasting ≥ 24 hours, and could be associated to seizure and/or focal neurological sign, in the absence of criteria for encephalitis criteria. We defined COVID-associated encephalopathy (CAE) if encephalopathy could not be accounted for by another cause such as toxic or metabolic factor according to the reporting clinician.
- Guillain-Barré Syndrome (GBS) was defined according to standard diagnosis criteria^7^.
- Acute meningitis was defined as meningeal syndrome without encephalitic course, and CSF WBC counts > 5/mm^3^.
- Patients with neurological manifestations that could not be classified into these five categories were classified as ‘others’

## RESULTS

The study was conducted in March-April 2020. Among the 259 CRF submitted, 37 patients were excluded for the following reasons: COVID-19 not confirmed (n=20), neurological symptoms non time-related to other symptoms of COVID-19 (n=12) or neurological symptoms linked to an exacerbation of chronic neurological disease (n=5). The population study (Figure 1) consisted of 222 COVID-19 patients with neurological manifestations from 46 centers in all the regions of continental France and overseas. Participant centers were university hospitals for 101/222 patients (45.5%) and non-university hospitals for 121/222 patients (54.5%). Physicians who filled out the forms were neurologists (146/222, 65.8%), infectious diseases or internal medicine specialists (43/222, 19.4%), intensivists (14/222, 6.3%), or other specialists (19/222, 8.6%). Most patients (217/222, 97.8%) required hospital admission.

**Figure 1.**
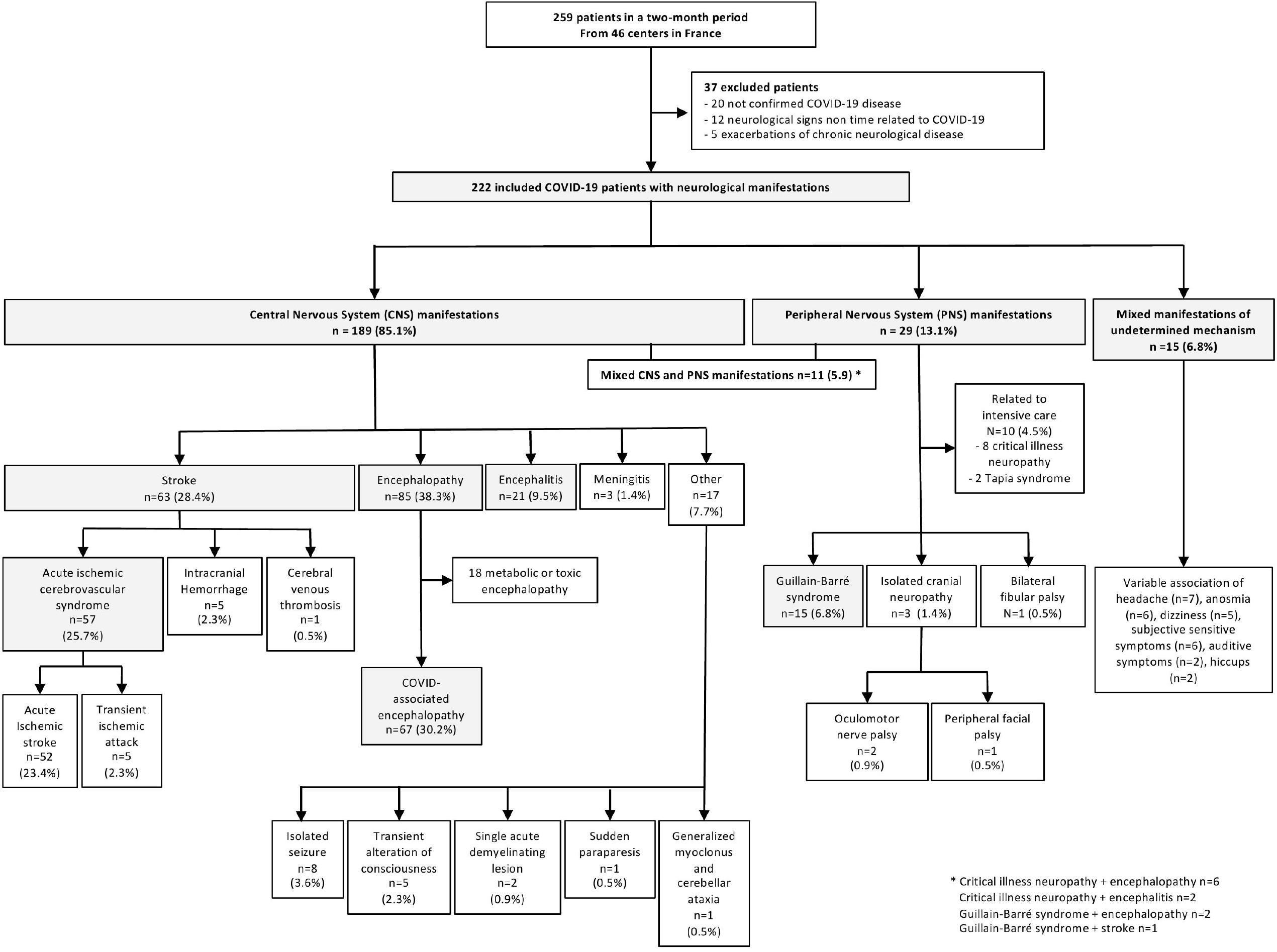
Study population of COVID-19 patients with neurological manifestations.

### General characteristics of COVID-19 patients with neurological manifestations

The general characteristics of the 222 patients are detailed in Table 1. Median age was 65 years (IQR 53-72), and 136 (61.3%) were male. Forty-seven patients (21.2%) had a past neurological history, mostly prior stroke (20, 9.0%) and neurodegenerative disease (17, 7.7%). The diagnosis of COVID-19 was confirmed by a positive SARS-CoV-2 nasopharyngeal rt-PCR in 86.5% (n=192) of the patients and by serology in 1.8% (n=4) of the patients. Twenty-six patients (11.7%) had a diagnosis on the basis of a typical clinical course and imagery. COVID-19 severity was mild or moderate in about half of the patients (120, 54.1%), severe or critical in the other half (102, 45.2%). The most commonly described neurological symptoms were altered mental status (117, 52.4%) and central focal deficit (97, 43.7%). Neurological work-up mostly included brain MRI (157, 70.7%) and CSF examination (97, 43.7%). SARS-CoV-2 PCR was performed in the CSF for 75 patients (33.8%) and was negative in 73/75 cases (97.3%). The median follow-up was 24 days (IQR 17-34). Twenty-eight patients (12.6%) died, mostly following acute respiratory distress syndrome (ARDS, n=17, 7.7%). Stroke was considered to be the cause of death in five patients (2.3%), following brain herniation due to extensive spontaneous intracerebral hematoma (n=2), severe hemorrhagic transformation after ischemic stroke (n=2) or extensive cerebral venous thrombosis (n=1).

**Table 1.**
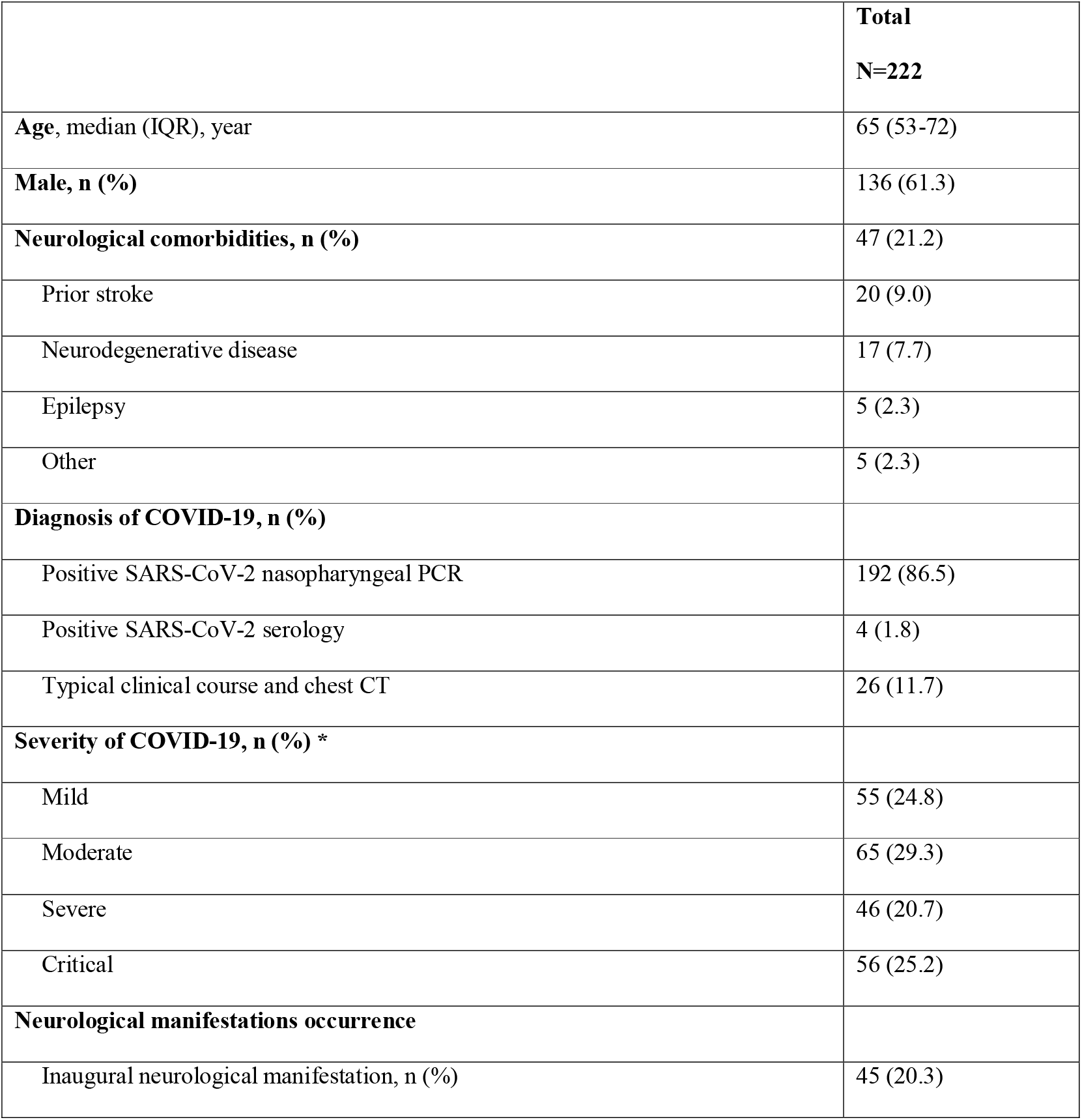
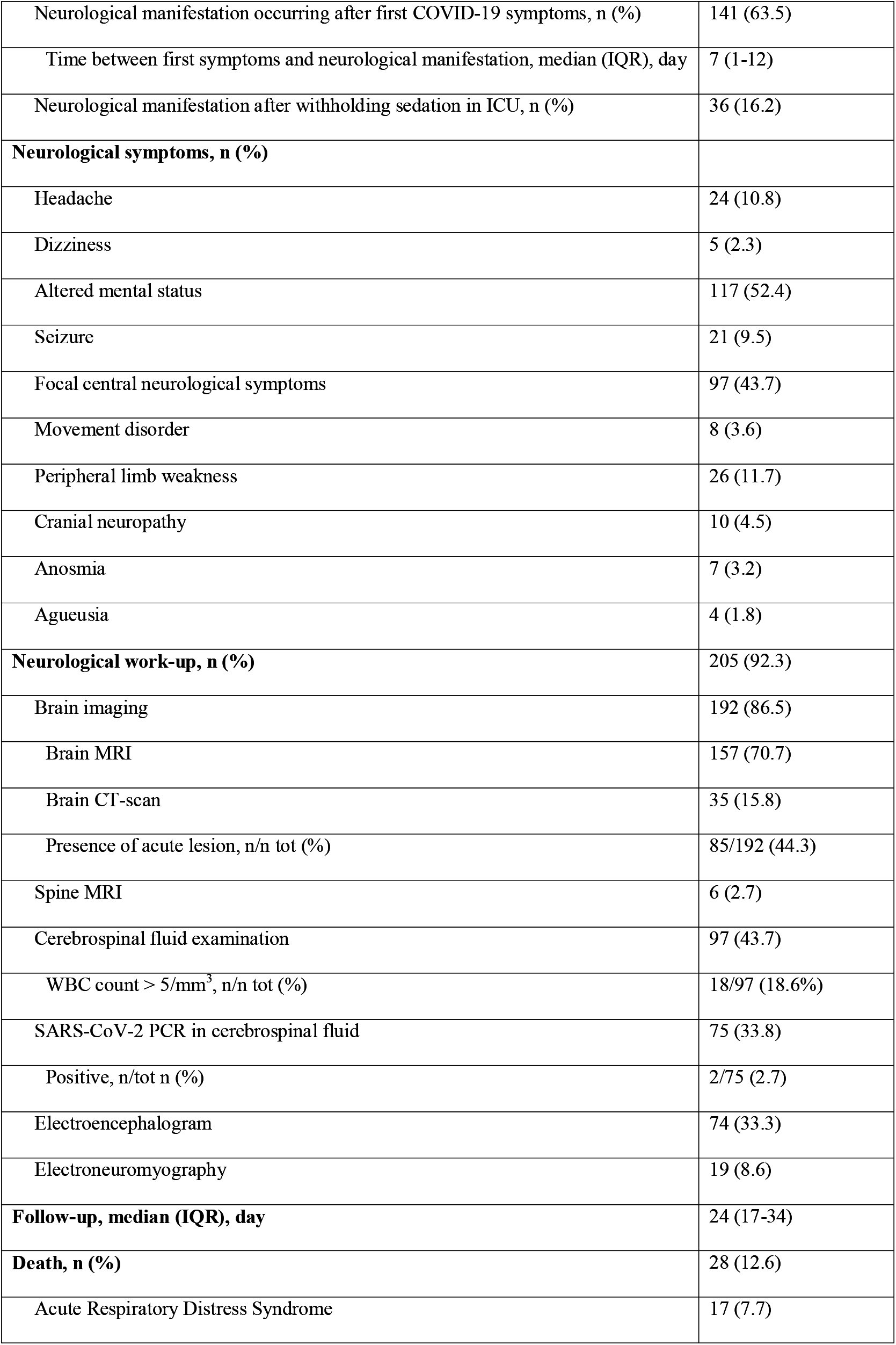
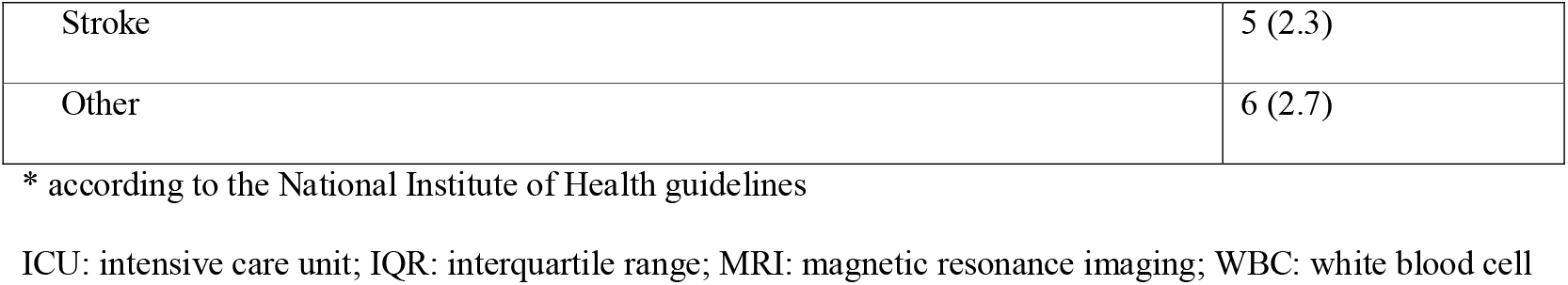
General characteristics of 222 COVID-19 patients with neurological manifestations.

### Clinical spectrum of neurological manifestations associated to SARS-CoV-2 infection

The spectrum of the neurological manifestations associated with SARS-CoV-2 infection is presented in Figure 1.

### CNS manifestations

One hundred eighty-nine patients (85.1%) had CNS manifestations, mostly encephalopathy (n=85, 38.3% of all patients), strokes (n=63, 28.4%), and encephalitis (n=21, 9.5%). The distribution of strokes was as follows: acute ischemic cerebrovascular syndromes (AICS, 57/63, 90.5% of all strokes) including 52 acute ischemic strokes and 5 transient ischemic attacks, intracranial hemorrhages (ICH, 5/63, 7.9%) and cerebral venous thromboses (CVT, 1/63, 1.6%). Among patients with encephalopathy, 67/85 (78%) were classified as COVID-associated encephalopathy (CAE). The 18 remaining patients had other factors accounting for encephalopathy: acute kidney injury in 10, medication in 6, complications of alcohol withdrawal in 4 patients. The other CNS manifestations were isolated seizures (n=8), transient loss of consciousness (n=5), meningitis (n=3), single acute demyelinating lesion (n=2), paraparesis (n=1) and generalized myoclonus with cerebellar ataxia (n=1).

### PNS manifestations

Twenty-nine patients (13.1%) had PNS manifestations, mostly Guillain-Barré syndrome (GBS, n=15, 6.8% of all patients). Ten other patients had peripheral complications of intensive care management, either critical illness neuropathy (n=8) or hypoglossal and pneumogastric nerves palsy following oro-tracheal intubation, known as Tapia syndrome (n=2). The remaining PNS manifestations were cranial neuropathy (n=3) and bilateral fibular nerve palsy (n=1). Eleven patients (5.9%) had mixed CNS and PNS manifestations, as detailed in Figure 1.

### Unclassified manifestations

Fifteen patients (6.8%) had mixed manifestations of undetermined mechanisms, including headache, dizziness, anosmia, auditory symptoms and subjective sensitive symptoms, in different combinations.

### Main neurological manifestations associated with COVID-19: AICS, encephalitis, CAE and GBS

AICS, encephalitis, CAE and GBS patients accounted for 71% (158/222) of the reported neurological manifestations in COVID-19 patients. Their characteristics are presented in table 2 and 3 and detailed below.

**Table 2.** Baseline and clinical characteristics of COVID-19 patients with acute ischemic cerebrovascular syndrome, encephalopathy, encephalitis, and Guillain-Barré Syndrome.

|  | **Acute ischemic**<br>**cerebrovascular**<br>**syndrome**<br>**(n=57)** | **Encephalitis**<br>**(n=21)** | **COVID**<br>**associated**<br>**encephalopathy**<br>**(n=67)** | **Guillain-**<br>**Barré**<br>**Syndrome**<br>**(n=15)** |
| --- | --- | --- | --- | --- |
| **Age**, median (IQR), year | 65 (55-78) | 67 (51-70) | 68 (61-75) | 59 (53-65) |
| **Male**, n (%) | 34 (59.6) | 15 (71.4) | 41 (60.3) | 13 (86.7) |
| **Medical history**, n (%) |  |  |  |  |
| Prior stroke, n (%) | 8 (14.0) | 0 (0) | 4 (6.0) | 1 (6.7) |
| Neurodegenerative disease, n (%) | 1 (1.8) | 1 (4.8) | 20 (29.9) | 0 (0) |
| Vascular comorbidities, n (%) * | 43 (75.4) | N/A* | N/A* | N/A* |
| **Severity of COVID-19**, n (%) |  |  |  |  |
| Mild | 21 (36.8) | 4 (19) | 6 (9.0) | 7 (46.7) |
| Moderate | 16 (28.1) | 7 (33.3) | 16 (23.9) | 3 (20.0) |
| Severe | 13 (22.8) | 3 (14.3) | 17 (25.4) | 1 (6.7) |
| Critical | 7 (12.3) | 7 (33.3) | 29 (43.3) | 4 (26.7) |
| **Neurological manifestations**<br>**occurrence** |  |  |  |  |
| Inaugural neurological<br>manifestation, n (%) | 14 (24.6) | 1 (4.8) | 15 (22.4) | 0 (0) |
| Neurological manifestation<br>occurring after first COVID-19<br>symptoms, n (%) | 40 (70.2) | 14 (66.7) | 32 (47.8) | 12 (80.0) |
| Time between first symptoms and neurological manifestation, median (IQR), day | 12 (7-18) | 7 (5-10) | 6 (3-8) | 18 (15-28) |
| Neurological manifestation after withholding sedation in ICU, n (%) | 3 (5.3) | 6 (28.6) | 20 (29.9) | 3 (20.0) |
| **Neurological symptoms, n (%)** |  |  |  |  |
| Headache | 2 (3.5) | 3 (14.3) | 6 (9.0) | 0 (0) |
| Altered mental status | 8 (14.0) | 21 (100) | 67 (100) | 3 (20.0) |
| Seizure | 1 (1.8) | 2 (9.5) | 7 (10.4) | 0 (0) |
| Focal central neurological symptoms | 56 (98.2) | 12 (57.1) | 13 (19.4) | 2 (13.3) |
| Motor or sensitive deficit | 42 (73.7%) | 2 (9.5) | 1 (1.5) | 1 (6.7) |
| Cerebellar ataxia | 6 (10.5%) | 6 (28.6) | 9 (13.4) | 0 (0) |
| Pyramidal syndrome | N/A | 6 (28.6) | 4 (6.0) | 0 (0) |
| Central oculomotor syndrome | 6 (10.5%) | 1 (4.8) | 1 (1.5) | 1 (6.7) |
| Movement disorder | 0 (0) | 6 (28.6) | 3 (4.5) | 1 (6.7) |
| Peripheral limb weakness | 1 (1.8) | 2 (9.5) | 7 (10.4) | 11 (73.3) |
| Cranial neuropathy | 0 (0) | 1 (4.8) | 2 (3.0) | 4 (26.7) |
| **Follow-up, median (IQR), day** | 24 (16-32) | 21 (18-29) | 28 (19-37) | 18 (14-29) |
| **Resolution of neurological symptoms, n (%)** | 21 (36.8) | 10 (47.6) | 34 (50.7) | 1 (6.7) |
| **Death, n (%)** | 9 (15.8) | 1 (4.8) | 10 (14.9) | 0 (0) |

\* Vascular comorbidities only collected for stroke patients. Data collected including hypertension, diabetes, obesity and cardiovascular diseases.
ICU: intensive care unit; IQR: interquartile range

**Table 3.**
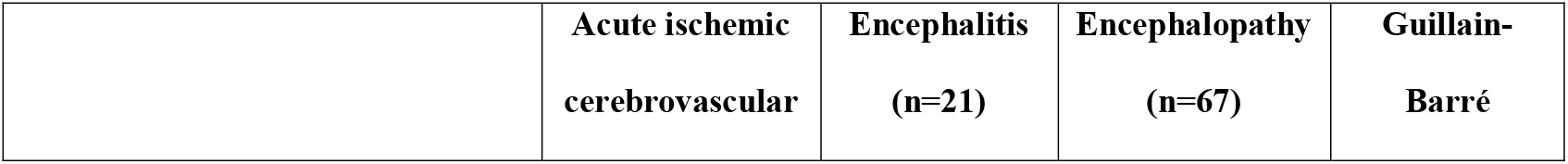
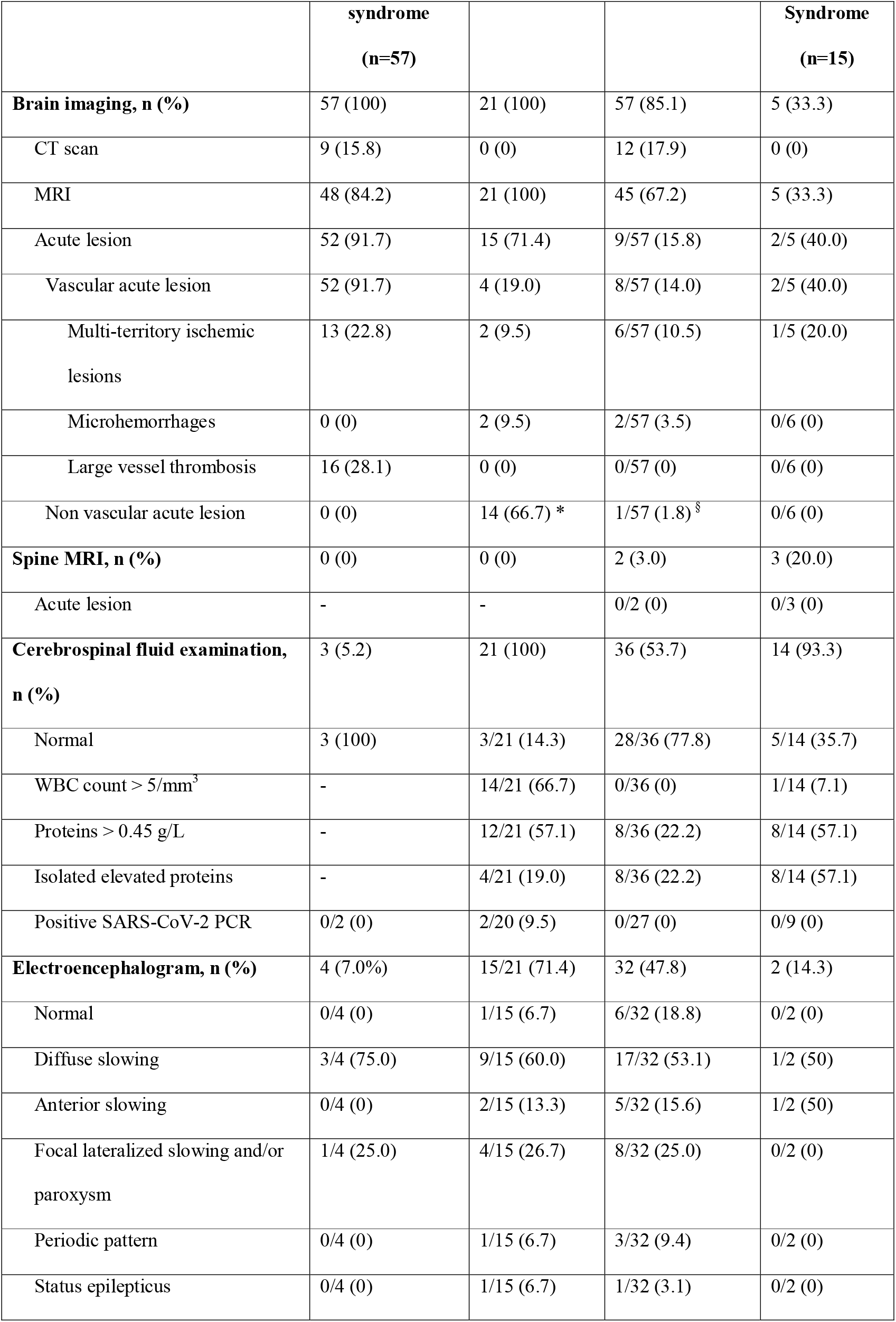
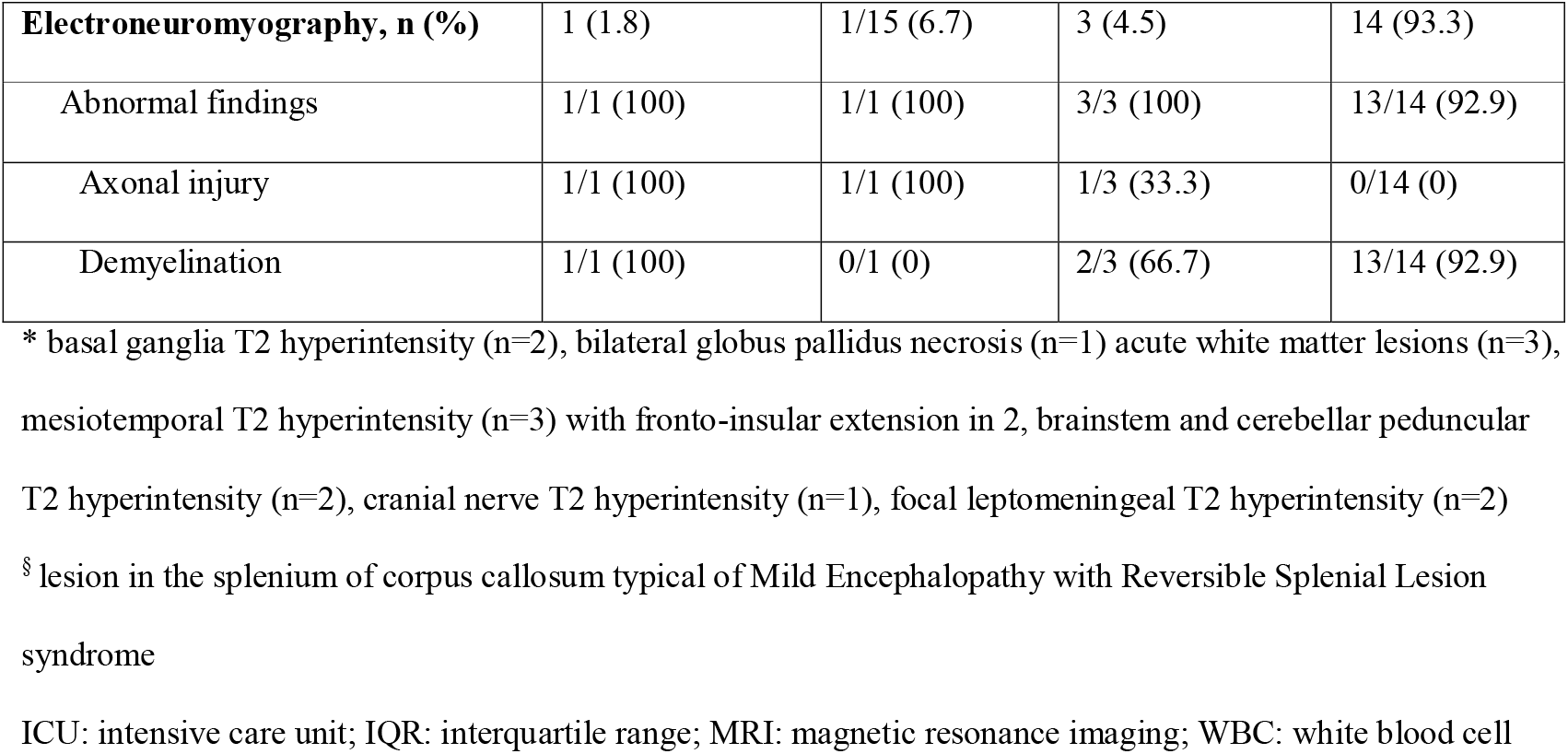
Neurological workup in COVID-19 patients with acute ischemic cerebrovascular syndrome, encephalitis, encephalopathy, and Guillain-Barré Syndrome.

|  | **Acute ischemic**<br>**cerebrovascular** | **Encephalitis**<br>**(n=21)** | **Encephalopathy**<br>**(n=67)** | **Guillain-**<br>**Barré** |
| --- | --- | --- | --- | --- |

|  | **syndrome**<br>**(n=57)** |  |  | **Syndrome**<br>**(n=15)** |
| --- | --- | --- | --- | --- |
| **Brain imaging, n (%)** | 57 (100) | 21 (100) | 57 (85.1) | 5 (33.3) |
| CT scan | 9 (15.8) | 0 (0) | 12 (17.9) | 0 (0) |
| MRI | 48 (84.2) | 21 (100) | 45 (67.2) | 5 (33.3) |
| Acute lesion | 52 (91.7) | 15 (71.4) | 9/57 (15.8) | 2/5 (40.0) |
| Vascular acute lesion | 52 (91.7) | 4 (19.0) | 8/57 (14.0) | 2/5 (40.0) |
| Multi-territory ischemic lesions | 13 (22.8) | 2 (9.5) | 6/57 (10.5) | 1/5 (20.0) |
| Microhemorrhages | 0 (0) | 2 (9.5) | 2/57 (3.5) | 0/6 (0) |
| Large vessel thrombosis | 16 (28.1) | 0 (0) | 0/57 (0) | 0/6 (0) |
| Non vascular acute lesion | 0 (0) | 14 (66.7) * | 1/57 (1.8) § | 0/6 (0) |
| **Spine MRI, n (%)** | 0 (0) | 0 (0) | 2 (3.0) | 3 (20.0) |
| Acute lesion | - | - | 0/2 (0) | 0/3 (0) |
| **Cerebrospinal fluid examination, n (%)** | 3 (5.2) | 21 (100) | 36 (53.7) | 14 (93.3) |
| Normal | 3 (100) | 3/21 (14.3) | 28/36 (77.8) | 5/14 (35.7) |
| WBC count > 5/mm<sup>3</sup> | - | 14/21 (66.7) | 0/36 (0) | 1/14 (7.1) |
| Proteins > 0.45 g/L | - | 12/21 (57.1) | 8/36 (22.2) | 8/14 (57.1) |
| Isolated elevated proteins | - | 4/21 (19.0) | 8/36 (22.2) | 8/14 (57.1) |
| Positive SARS-CoV-2 PCR | 0/2 (0) | 2/20 (9.5) | 0/27 (0) | 0/9 (0) |
| **Electroencephalogram, n (%)** | 4 (7.0%) | 15/21 (71.4) | 32 (47.8) | 2 (14.3) |
| Normal | 0/4 (0) | 1/15 (6.7) | 6/32 (18.8) | 0/2 (0) |
| Diffuse slowing | 3/4 (75.0) | 9/15 (60.0) | 17/32 (53.1) | 1/2 (50) |
| Anterior slowing | 0/4 (0) | 2/15 (13.3) | 5/32 (15.6) | 1/2 (50) |
| Focal lateralized slowing and/or paroxysm | 1/4 (25.0) | 4/15 (26.7) | 8/32 (25.0) | 0/2 (0) |
| Periodic pattern | 0/4 (0) | 1/15 (6.7) | 3/32 (9.4) | 0/2 (0) |
| Status epilepticus | 0/4 (0) | 1/15 (6.7) | 1/32 (3.1) | 0/2 (0) |
| **Electroneuromyography, n (%)** | 1 (1.8) | 1/15 (6.7) | 3 (4.5) | 14 (93.3) |
| Abnormal findings | 1/1 (100) | 1/1 (100) | 3/3 (100) | 13/14 (92.9) |
| Axonal injury | 1/1 (100) | 1/1 (100) | 1/3 (33.3) | 0/14 (0) |
| Demyelination | 1/1 (100) | 0/1 (0) | 2/3 (66.7) | 13/14 (92.9) |

\* basal ganglia T2 hyperintensity (n=2), bilateral globus pallidus necrosis (n=1) acute white matter lesions (n=3), mesiotemporal T2 hyperintensity (n=3) with fronto-insular extension in 2, brainstem and cerebellar peduncular T2 hyperintensity (n=2), cranial nerve T2 hyperintensity (n=1), focal leptomeningeal T2 hyperintensity (n=2)
§ lesion in the splenium of corpus callosum typical of Mild Encephalopathy with Reversible Splenial Lesion syndrome
ICU: intensive care unit; IQR: interquartile range; MRI: magnetic resonance imaging; WBC: white blood cell

#### Baseline characteristics (table 2)

Median age was 65 years (IQR 55-78) in AICS patients, 67 years (IQR 51-70) in encephalitis patients, 68 years (IQR 61-75) in CAE patients and 59 years (IQR 53-65) in GBS patients. The proportion of men was higher in each group, particularly in encephalitis patients (15/21, 71.4%) and GBS patients (13/15, 86.7%). Concerning medical history, 20/67 (29.9%) of CAE patients had neurodegenerative disease, and 43/57 (75.4%) of AICS patients had vascular comorbidities, including 8/52 (14.0%) with prior stroke.

The majority of CAE patients suffered from severe to critical COVID-19 illness (46/67, 68.7%). In GBS and AICS patients, mild and moderate COVID-19 illness predominated, respectively 10/15 (66.7%) and 37/57 (64.9%). AICS was inaugural of COVID-19 in 14/57 patients (24.6%), and CAE in 15/67 patients (22.4%). Neurological symptoms were first observed when sedation was withheld for 20/67 CAE patients (29.9%), 6/21 encephalitis patients (28.6%), and 3/15 GBS patients (20.0%). The remaining patients had neurological manifestations several days after the first COVID-19 symptoms, with a median delay of 6 (IQR 3-8) and 7 (IQR 5-10) days respectively in CAE and encephalitis patients, 12 days (IQR 7-18) in AICS and 18 days (IQR 15-28) in GBS.

#### Clinical course (table 2)

Regarding AICS, focal central neurological symptoms were present in 56/57 patients (98.2%), and altered mental status in 8/57 (14.0%). All patients with encephalitis or CAE had altered mental status. Twelve out of 21 encephalitis patients (57.1%) had focal central neurological symptoms, mostly cerebellar ataxia and pyramidal syndrome, and six (28.6%) had movement disorders, mostly tremor and myoclonus. Focal neurological symptoms were also present in 13/67 CAE patients (19.4%) mostly cerebellar ataxia. Seizure rate was approximately the same in encephalitis (2/21, 9.5%) and in CAE (7/67, 10.4%). Eleven out of 15 patients (73.3%) with GBS had progressive weakness in both arms and legs that could be associated with sensory symptoms. Among these 11 patients, one had oculomotor nerve palsy and another had bilateral facial nerve palsy. Three other patients (20.0%) had proprioceptive sensory loss without limb weakness, associated with facial palsy in one. One patient (6.7%) had bilateral facial palsy and areflexia.

#### Neurological work-up (table 3)

Large vessel thrombosis was observed in 16/57 patients with AICS (28.1%), located in internal carotid artery (n=9) and/or the proximal segment of the middle cerebral artery (n=6), or in basilar artery (n=1). Thirteen patients (22.8%) had multi territory ischemic strokes. AICS was cryptogenic in 38/57 patients (66.7%). Etiological workup highlighted 9 embolic heart disease (atrial fibrillation in 4, paradoxical embolism in 2, valvular heart disease in 2, myocarditis in 1), 7 atheromatous stenosis, 1 cerebral small vessel disease, 1 intracranial vasculopathy and 1 carotid artery dissection. In encephalitis patients, CSF examination demonstrated lymphocytic pleiocytosis from 6 to 77 WBC/mm^3^ in 14/21 patients (66.7%) and positive SARS-CoV-2 PCR in 2/20 (9.5%). Brain MRI showed heterogeneous acute non vascular lesion in 14/21 patients (66.7%), listed in table 3. Four patients (19.0%) also had small ischemic lesion or microhemorrhages. Regarding CAE patients, 57/67 had brain imaging (85.1%) showing nine with acute lesions, mostly small ischemic ones (table 3). Electroencephalogram was performed in 15/21 encephalitis patients (71.4%) and 32/67 CAE patients (47.8%). The proportion of the different patterns among the two groups was very close (table 3) with 55% of non-specific diffuse slowing (n=26/47), 15% of anterior slowing (n=7/47), 26% of focal lateralized abnormalities (n=12/47) and 9% of periodic pattern (n=4/47). In patients with GBS, CSF examination demonstrated isolated elevated proteins in 8/14 patients (57.1%), ranging from 0.49 to 2.36 g/L, and negative SARS-CoV-2 PCR in the 9 patients tested. One patient had mild lymphocytic pleiocytosis with 12 WBC/mm^3^. Electroneuromyography was abnormal in 13/14 patients (92.9), suggestive of demyelination in all of them.

#### Outcome (table 2)

Regarding encephalitis, 10/21 patients (47.6%) fully recovered, 3 of whom received corticosteroids (CS). In patients with persistent neurological symptoms (10/21, 47.6%) one received CS and another intravenous immunoglobulins (IVIg). Out of 67 patients with CAE, 34 (50.7%) recovered spontaneously. Two patients received corticosteroids with partial improvement. Most patients with GBS were treated with IVIg for 14/15 (93.3%). Two of them required mechanical ventilation. The mortality rate during follow-up was 15.8% in AICS patients, 14.9% in CAE patients, 4.8% in encephalitis patients, 0% in GBS patients. To note, among the 18 patients with toxic metabolic encephalopathy who had been excluded from the CAE group, five deaths occurred (27.8%).

### Other neurological manifestations associated with COVID-19

Fifty patients had neurological manifestations that did not fulfill criteria for AICS, encephalitis, CAE or GBS, nor were they related to intensive care complications or metabolic or toxic encephalopathy. These neurological manifestations were very heterogeneous (Figure 1, Table 4).

**Table 4.**
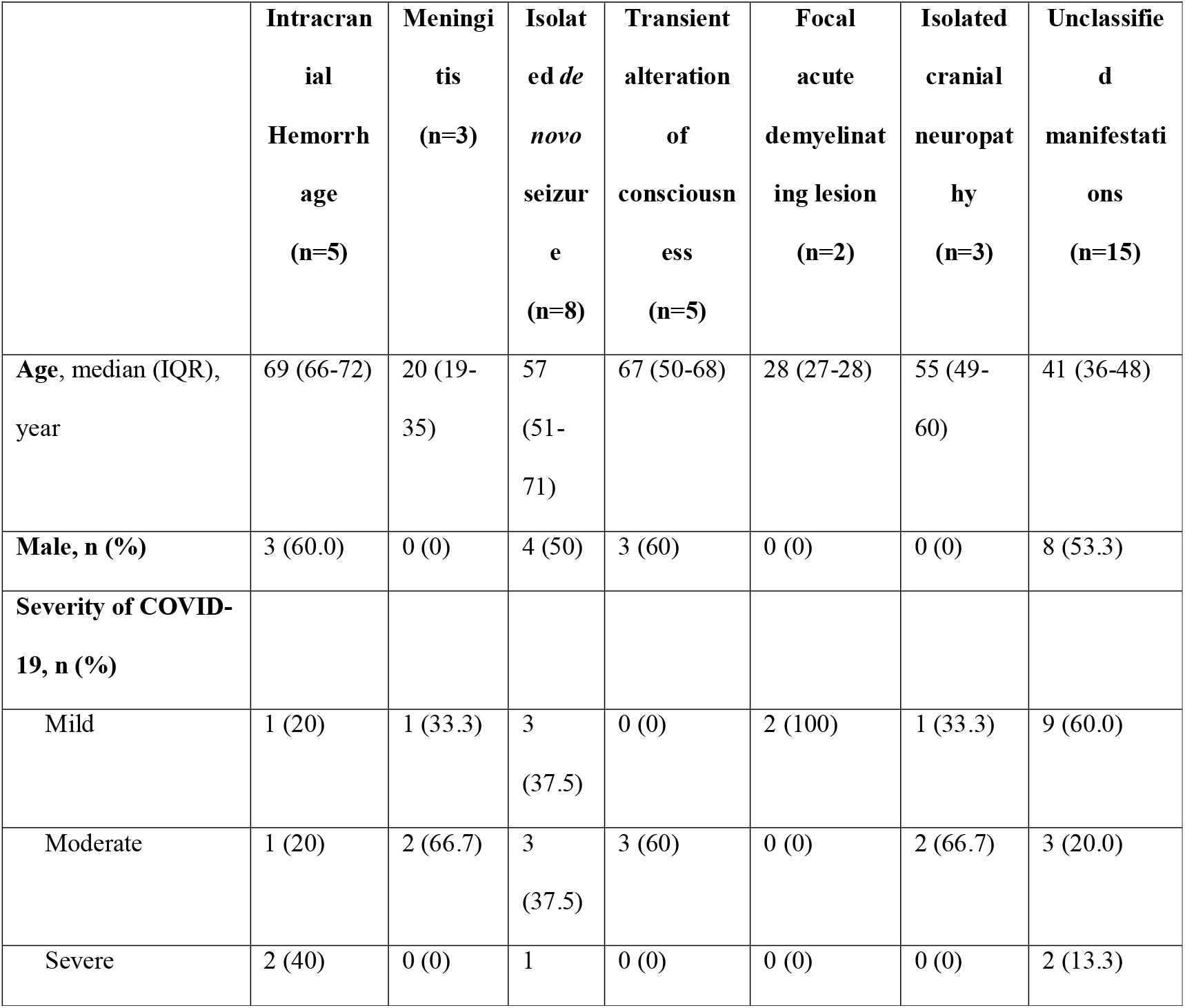
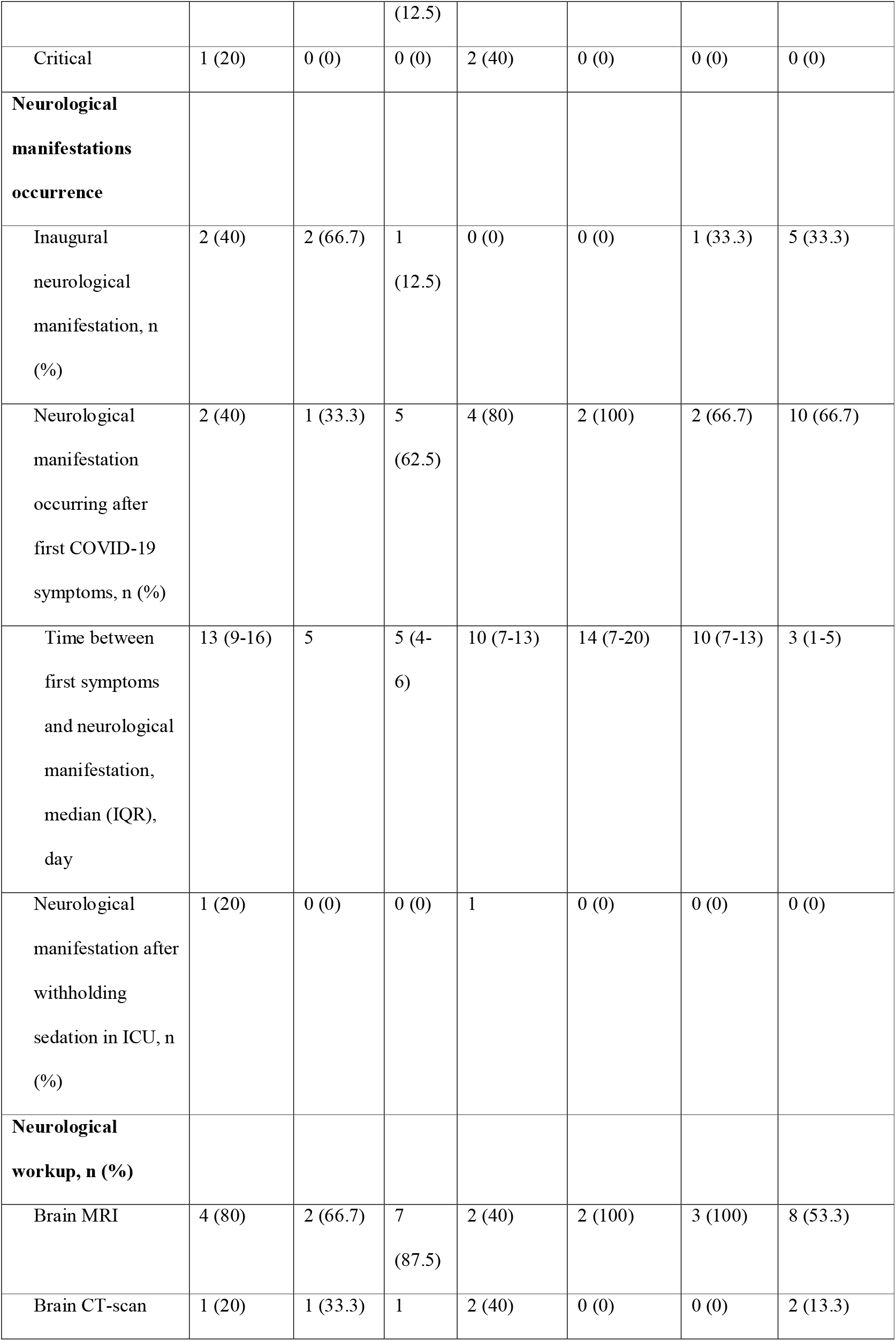
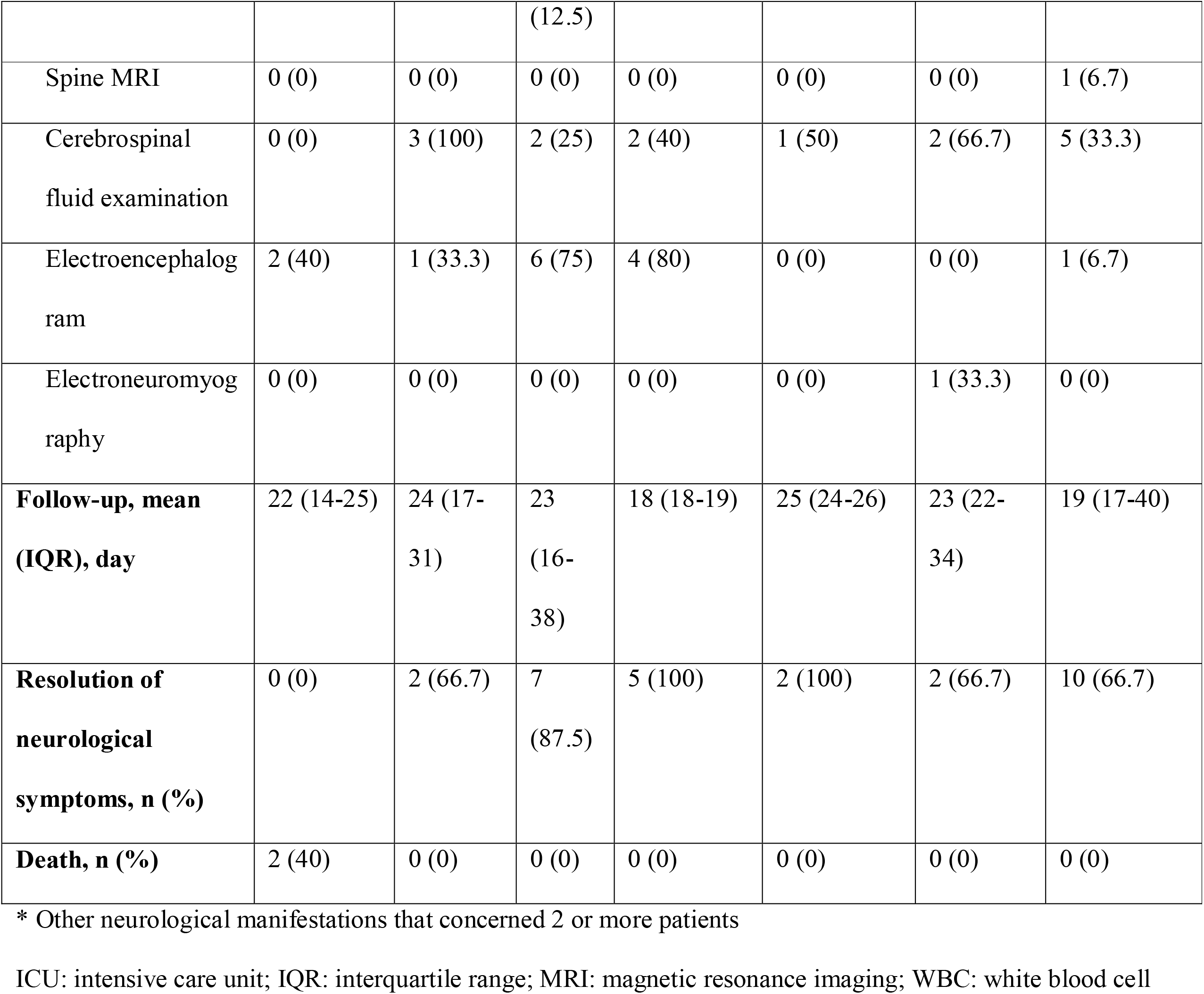
Characteristics of COVID-19 patients with other neurological manifestations*.

#### Acute benign lymphocytic meningitis

Three young patients, with a median age of 20 years (IQR 19-35), had acute benign lymphocytic meningitis in a context of mild or moderate COVID-19 disease. CSF examination demonstrated lymphocytic pleiocytosis from 12 to 102 WBC/mm^3^, a normal protein level and negative SARS-CoV-2 PCR in the two samples that were tested.

#### Non ischemic strokes

Five patients had ICH. Three of them had deep ICH with a history of chronic arterial hypertension. Another patient had lobar ICH with no evidence of an underlying vascular malformation or tumor after extensive work-up including brain MRI and cerebral angiography. The last patient had multiple lobar ICH in a context of severe COVID-19 disease with acute kidney injury and hemolytic anemia. Two out of the five patients with ICH (40%) died. One patient had CVT revealed by a large parenchymal hematoma and a fulminant intracranial hypertension, causing death within a few days.

#### Peripheral neuropathy

Two patients had oculomotor nerve palsy and one had facial peripheral nerve palsy with spontaneous clinical improvement in most cases (2/3, 66.7%) and no evidence for an underlying acute inflammatory demyelinating neuropathy. One patient had bilateral fibular palsy with an asymmetrical motor predominant neuropathy of undetermined mechanism.

#### Isolated seizure and transient alteration of consciousness

Eight patients had isolated de novo seizure and five had transient alteration of consciousness, with resolution of symptoms in less than 24 hours and no other encephalopathic features. Semeiology of seizure were as follows: generalized tonic-clonic seizure (n=6), focal motor seizure (n=1), focal non-motor seizure with chewing movement (n=1). One patient experienced recurrence of generalized seizure after 24 hours and required long term antiepileptic medication, with etiological negative work-up.

#### Focal acute demyelinating lesion

Two young women with no medical history revealed multiple sclerosis with an acute demyelinating MRI lesion, optic neuritis in one and cerebellar peduncle active lesion in one, with several old lesions.

#### Other rare neurological manifestations

One patient had paraparesis of undetermined mechanism, with no evidence for myelitis on spine MRI and no evidence for Guillain-Barré syndrome on CSF examination and electroneuromyography. Another patient had generalized myoclonus with cerebellar ataxia. Brain MRI revealed no significant abnormalities and CSF examination was not available. He improved partially without any treatment.

#### Mixed symptoms of unclassified mechanisms

Fifteen patients (6.8%) had different combinations of nonspecific symptoms such as headache or dizziness, that could be associated with anosmia or hiccups. Most patients (12/15, 80%) had mild or moderate COVID-19 disease. Neurological symptoms appeared early in the evolution of the disease as they were inaugural for 5 patients and appeared in a median delay of 3 days for the others.

## DISCUSSION

To our knowledge, this is one of the largest series of COVID-19 patients with neurological manifestations. It provides detailed data about clinical and diagnostic aspects including short-term prognosis. Our results highlighted the broad spectrum of neurological manifestations associated with SARS-CoV-2 infection, and identified some major trends. The majority of neurological manifestations were CAE (67/222, 30%), AICS (57/222, 26%), encephalitis (21/222, 10%), and GBS (15/222, 7%). Neurological manifestations appeared after first COVID-19 symptoms with a median delay of 6 days in CAE, 7 days in encephalitis, 12 days in AICS and 18 days in GBS. Around 25% of CAE and AICS were inaugural and 30% of CAE and encephalitis were diagnosed after discontinuation of sedation. A combination of nonspecific symptoms such as headache or dizziness, not related to another neurological disease, were reported in 7% of the patients (15/222) and appeared early in the course of COVID-19. With a median delay of follow-up of 24 days, our registry highlighted a high rate of short-term mortality in COVID-19 patients with CAE and AICS, around 15% (19/124). The other reported fatal cases concerned five patients with metabolic-toxic encephalopathy, two patients with ICH, one patient with CVT and one patient with encephalitis.

Altered mental status was the most common neurological symptom reported in 52% of patients in our registry. Several cohorts of hospitalized patients with COVID-19 have shown a significant proportion of impaired consciousness on admission, ranging from 7.5 to 20% ^1,3,4,8^. The classification and description of these patients in our registry highlight different possible pathogenic pathways. Indeed, patients with altered mental status may have encephalitis, in presence of meningitis and/or a compatible lesion on cerebral MRI. Encephalitis represented up to 10% of patients in our registry and they were more likely to have focal neurological signs and abnormal movements compared to patients with CAE. The aspects of brain MRI were very heterogeneous, consistent with the few published cases of encephalitis: white matter lesion and / or basal ganglia and thalami involvement suggestive of acute disseminated encephalomyelitis^9^ or acute necrotizing encephalopathy^10–12^, other nonspecific diffuse involvement of white matter^13,14^, mesio-temporal lesion^8,15^ with possible frontoinsular extension, leptomeningeal abnormalities^4^ and brainstem lesions^16^. Cases with unremarkable MRI have also been published^17,18^. Only two patients in our registry had a positive SARS-CoV-2 PCR in the CSF. In line with our data, only two encephalitis with positive SARS-CoV-2 PCR in CSF have been reported by the time of writing^9,15^. In our series the short-term outcome was generally favorable without any specific treatment. All of these elements suggest rather a para-infectious mechanism than a direct neuropathogenicity of the virus, but neuropathological studies remain scarce. In a recent autopsy study of six patients who died from COVID-19, Von Weyhern and al^19^ highlighted the presence of lymphocytic panencephalitis and meningitis, but whether the observed lesions were a direct consequence of virus infiltration or resulted from an immune response could not be established. Another published case documented the presence of SARS-CoV-2 in endothelial and neural cells of frontal lobe sections^20^ in a patient with negative SARS CoV 2 PCR in postmortem CSF sample. This raises the question of the sensitivity of SARS-CoV-2 PCR testing in CSF, currently unknown. In addition, twenty percent of patients with encephalitis in our series demonstrated microvascular lesions on brain MRI, suggesting potential implication of a COVID-19 associated endotheliitis ^21,22^ or coagulopathy^23^. In the autopsy study of Von Weyhern and al^19^, diffuse petechial brain hemorrhages were associated with encephalitic pathological features, with no conspicuous endotheliitis.

Another autopsy study^24^ demonstrated the presence of acute hypoxic injury in the cerebrum and cerebellum with no thrombi, encephalitis or vasculitis, and no cytoplasmic viral staining. This study concerned 18 patients having encephalopathic features associated with fatal COVID-19 disease. In our registry, we described a group of 67 patients with CAE, defined by persistent confusional state without encephalitis criteria or obvious toxic-metabolic coexisting factor. CAE patients had more severe forms of COVID-19, as previously shown in other studies^3,8^, with one third of them exhibiting delirium after intensive care management when sedation was withdrawn. Helms et al described the same course in 67% of patients with COVID-19 related ARDS^4^. We can therefore hypothesize that COVID-induced sepsis^25^ may lead to a septic-associated encephalopathy (SAE). There are indeed many points suggestive of SAE in our CAE patients: age, previous cognitive impairment, illness severity, focal deficit and seizures, tremor, myoclonus and acute vascular lesion on brain MRI^26^. SAE can also induce posterior reversible encephalopathy syndrome^27^ which has not been reported in our registry but some cases have been published in COVID-19 patients^8,28^. The release of pro-inflammatory cytokines is one of the key pathogenic pathways suggested in SAE, and is thought to play a central role in severe COVID-19^29^. The cytokine storm is also one of the possible pathogenic pathways in mild encephalopathy with reversible splenial lesion syndrome, that is described in one patient in our registry and in one patient in the literature^30^.

In patients with encephalitis and CAE, electroencephalographic features were relatively close and mainly consisted of a nonspecific generalized background slowing, indicating diffuse cerebral dysfunction. Some published cases suggested a more specific pattern in COVID-19 patients, showing in frontal areas the presence of monomorphic biphasic delta slow waves with a periodic organization ^31,32^. Our study found only 15% of anterior slowing and 9% of periodic pattern in patients with encephalitis and CAE.

AICS was the second most common manifestation in our series, accounting for 26% of patients. Although 75% of AICS patients had vascular comorbidities, our study highlights some features for which there is growing evidence in other published series: high prevalence of large vessels stroke^33,34^, multi-territory involvement^34^, undetermined etiology^35^, and high mortality rate^35^. COVID-related coagulopathy^23^ probably plays a decisive role in these thrombotic cryptogenic ischemic strokes. AICS was not associated with particularly severe manifestations of COVID-19 in our series, contrary to what Mao et al^3^ had suggested. GBS represented approximately 7% of patients in the registry, and occurred with a median delay of 18 days after the first manifestations of COVID-19. Several cases and case series are currently reported in the literature^36–41^ and an Italian study has demonstrated that the incidence of GBS increased in their region during the COVID-19 epidemics compared with the three previous years^42^. No Miller Fisher syndrome case has been reported in our registry, and published cases are few and far between’^43,44^.

Patients with mixed manifestations of undetermined mechanisms such as headache, dizziness, subjective sensitive signs and anosmia have been reported to our registry in about 7% of patients. Headache and dizziness are frequently reported in cohorts of COVID-19 patients, ranging from 8 to 14% and 6 to 17% respectively^1,3,8^. Anosmia was present in 3% of our patients and in about 5% in other series of hospitalized COVID-19 patients^3,8^ whereas its prevalence reached 86% in a dedicated European multicentric study^45^. In our cohort that mainly involved hospitalized patients, these symptoms may have been underreported.

The other neurological manifestations reported in our registry are much rarer in occurrence and would require larger studies to be appropriately described. We report three cases of acute benign lymphocytic meningitis, which has not been reported in association with COVID-19 to date. We also report two patients with Tapia syndrome, a rare complication of orotracheal intubation, with one similar case recently published ^46^.

Our study has some limitations. First, this is a retrospective registry with inherent reporting biases, which should lead us to interpret with caution the different proportions of neurological manifestations. Headache, dizziness and anosmia were not particularly frequent in our registry, probably underreported and/or associated with milder forms of COVID-19 disease. Secondly, the data are entirely descriptive and based on the report at a definite time period during the French outbreak. We used a deliberately simplified CRF given the exceptional working load of medical teams, whose first mission during the outbreak was the management of patients. There was therefore no exhaustive collection of medical history other than neurological comorbidities and vascular comorbidities for AICS, nor of biological parameters. The main objective was to provide a wide overview of neurological manifestations associated with SARS-CoV-2 infection. The analysis of the mechanisms and potential risk factors for nervous system involvement in COVID-19 is therefore limited. Further studies are necessary to clarify these points.

## CONCLUSION

Our study highlights the broad spectrum of neurological manifestations associated with SARS-CoV-2 infection, probably related to different pathogenic pathways. Although encephalopathies were the most frequently reported manifestation, possibly linked to sepsis and cytokine storm, encephalitis was described in 10% of cases. However, we found few arguments to support the hypothesis of direct SARS-CoV-2 neuropathogenicity: a large majority of SARS-CoV-2 PCR in cerebrospinal fluid (73/75, 97.3%) were negative, and the short-term outcome of patients with encephalitis was generally favorable. Ischemic strokes were also frequently reported, as well as GBS, which occurred later in the course of the disease (18 days as compared to 7 days for encephalitis or 12 days for strokes). Other rare neurological manifestations such as meningitis or cranial neuropathy require comparison with other studies and registries to understand their potential link with SARS-CoV-2 infection.

## Data Availability

The data that support the findings of this study are available on request from the corresponding author, EM. The data are not publicly available due to information that could compromise the privacy of research participants.

## ACKNOWLEDGEMENTS

We acknowledge the support of the National College of General Hospital Neurologists (CNNHG), the French Society for Infectious Diseases (SPILF), the Association of French-Speaking Liberal Neurologists (ANLLF), the Multiple Sclerosis French Society (SF-SEP), REACTing, a French multi-disciplinary collaborative network working on emerging infectious diseases, and the CoCo Neurosciences Study Group.

We acknowledge the support of Dr. Jérôme Aboab and the Clinical Research Unit of Saint-Denis for obtaining the approval of the national ethics committees.

Preliminary results of this work have been presented at the European Academy of Neurology (EAN) in May, 2020, by Pr. Elena MORO.

## AUTHOR CONTRIBUTIONS

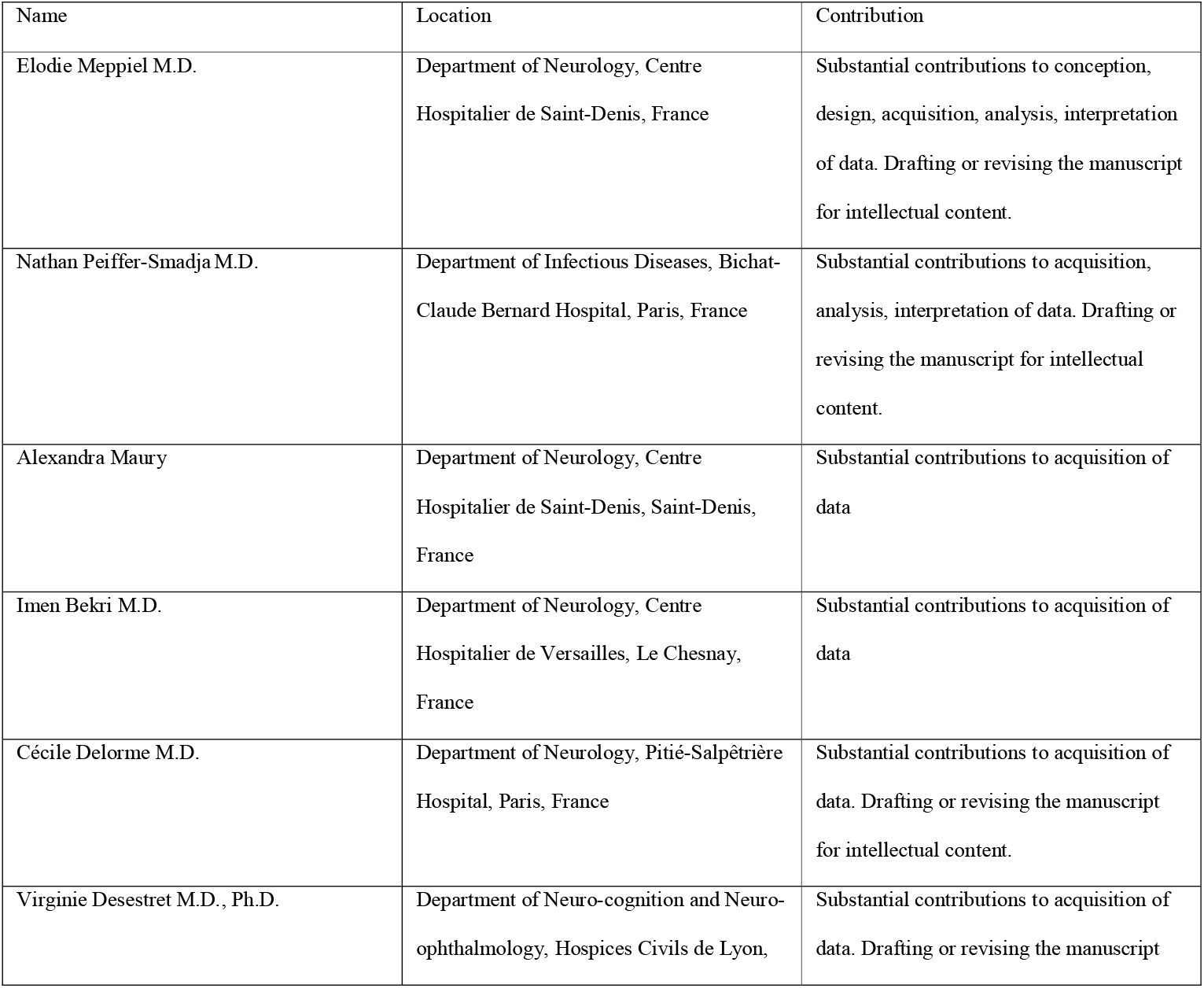
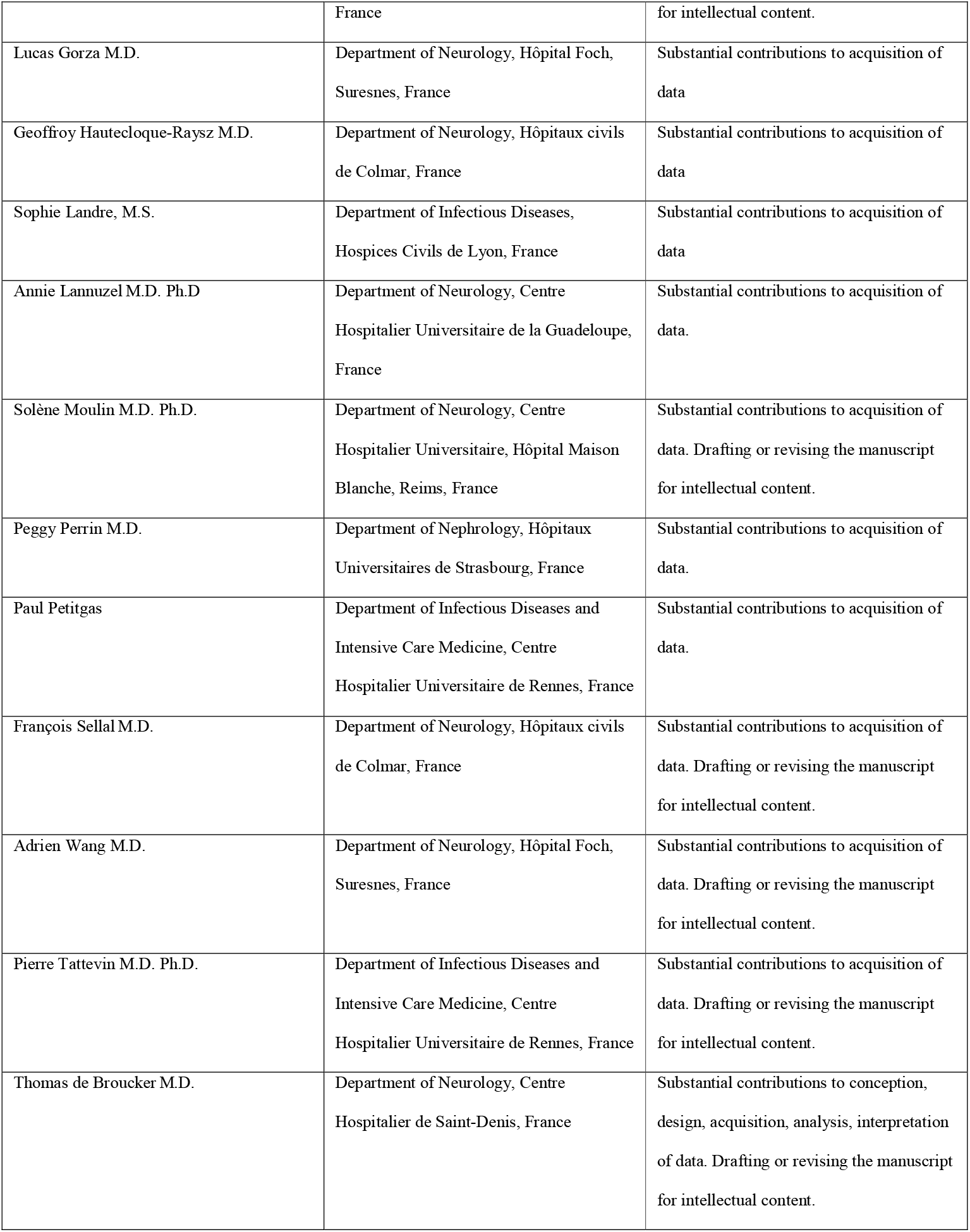

---

**Supplementary Appendix 1: contributors to the NeuroCOVID registry**

## REFERENCES

1. Chen N, Zhou M, Dong X, et al. Epidemiological and clinical characteristics of 99 cases of 2019 novel coronavirus pneumonia in Wuhan, China: a descriptive study. The Lancet 2020;395(10223):507–13.

2. Goyal P, Choi JJ, Pinheiro LC, et al. Clinical Characteristics of Covid-19 in New York City. New England Journal of Medicine 2020;0(0):null.

3. Mao L, Jin H, Wang M, et al. Neurologic Manifestations of Hospitalized Patients With Coronavirus Disease 2019 in Wuhan, China. JAMA Neurol [Internet] 2020 [cited 2020 May 6];Available from: https://jamanetwork.com/journals/jamaneurology/fullarticle/2764549

4. Helms J, Kremer S, Merdji H, et al. Neurologic Features in Severe SARS-CoV-2 Infection. N Engl J Med 2020;NEJMc2008597.

5. Zubair AS, McAlpine LS, Gardin T, Farhadian S, Kuruvilla DE, Spudich S. Neuropathogenesis and Neurologic Manifestations of the Coronaviruses in the Age of Coronavirus Disease 2019: A Review. JAMA Neurol [Internet] 2020 [cited 2020 Jun 1];Available from: https://jamanetwork.com/journals/jamaneurology/fullarticle/2766766

6. Management of COVID-19 | Coronavirus Disease COVID-19 [Internet]. COVID-19 Treatment Guidelines. [cited 2020 May 17];Available from: https://www.covid19treatmentguidelines.nih.gov/overview/management-of-covid-19/

7. Asbury AK, Cornblath DR. Assessment of current diagnostic criteria for Guillain-Barr□ syndrome. Ann Neurol 1990;27(S1):S21–4.

8. Romero-Sánchez CM, Díaz-Maroto I, Fernández-Díaz E, et al. Neurologic manifestations in hospitalized patients with COVID-19: The ALBACOVID registry. Neurology 2020;10.1212/WNL.0000000000009937.

9. Novi G, Rossi T, Pedemonte E, et al. Acute disseminated encephalomyelitis after SARS-CoV-2 infection. Neurol Neuroimmunol Neuroinflamm 2020;7(5):e797.

10. Poyiadji N, Shahin G, Noujaim D, Stone M, Patel S, Griffith B. COVID-19–associated Acute Hemorrhagic Necrotizing Encephalopathy: CT and MRI Features. Radiology 2020;201187.

11. Dixon L, Varley J, Gontsarova A, et al. COVID-19-related acute necrotizing encephalopathy with brain stem involvement in a patient with aplastic anemia. Neurol Neuroimmunol Neuroinflamm 2020;7(5):e789.

12. Virhammar J, Kumlien E, Fällmar D, et al. Acute necrotizing encephalopathy with SARS-CoV-2 RNA confirmed in cerebrospinal fluid. Neurology 2020;10.1212/WNL.0000000000010250.

13. Kandemirli SG, Dogan L, Sarikaya ZT, et al. Brain MRI Findings in Patients in the Intensive Care Unit with COVID-19 Infection. Radiology 2020;201697.

14. Brun G, Hak J-F, Coze S, et al. COVID-19—White matter and globus pallidum lesions: Demyelination or small-vessel vasculitis? Neurol Neuroimmunol Neuroinflamm 2020;7(4):e777.

15. Moriguchi T, Harii N, Goto J, et al. A first case of meningitis/encephalitis associated with SARS-Coronavirus-2. International Journal of Infectious Diseases 2020;94:55–8.

16. Wong PF, Craik S, Newman P, et al. Lessons of the month 1: A case of rhombencephalitis as a rare complication of acute COVID-19 infection. Clin Med (Lond) 2020;

17. Bernard-Valnet R, Pizzarotti B, Anichini A, et al. Two patients with acute meningo-encephalitis concomitant to SARS-CoV-2 infection. Eur J Neurol 2020;

18. Pilotto A, Odolini S, Masciocchi S, et al. Steroid-Responsive Encephalitis in Coronavirus Disease 2019. Annals of Neurology [Internet] [cited 2020 Jun 29];n/a(n/a). Available from: https://onlinelibrary.wiley.com/doi/abs/10.1002/ana.25783

19. von Weyhern CH, Kaufmann I, Neff F, Kremer M. Early evidence of pronounced brain involvement in fatal COVID-19 outcomes. The Lancet 2020;S0140673620312824.

20. Paniz□Mondolfi A, Bryce C, Grimes Z, et al. Central nervous system involvement by severe acute respiratory syndrome coronavirus-2 (SARS-CoV-2). Journal of Medical Virology 2020;92(7):699–702.

21. Varga Z, Flammer AJ, Steiger P, et al. Endothelial cell infection and endotheliitis in COVID-19. Lancet 2020;395(10234):1417–8.

22. Hanafi R, Outteryck O. COVID-19 Neurologic Complication with CNS Vasculitis-Like Pattern. :4.

23. Tang N, Li D, Wang X, Sun Z. Abnormal coagulation parameters are associated with poor prognosis in patients with novel coronavirus pneumonia. J Thromb Haemost 2020;18(4):844–7.

24. Solomon IH, Normandin E, Bhattacharyya S, et al. Neuropathological Features of Covid-19. N Engl J Med 2020;NEJMc2019373.

25. Li H, Liu L, Zhang D, et al. SARS-CoV-2 and viral sepsis: observations and hypotheses. The Lancet 2020;395(10235):1517–20.

26. Mazeraud A, Righy C, Bouchereau E, Benghanem S, Bozza FA, Sharshar T. Septic-Associated Encephalopathy: a Comprehensive Review. Neurotherapeutics [Internet] 2020 [cited 2020 May 23];Available from: http://link.springer.com/10.1007/s13311-020-00862-1

27. Racchiusa S, Mormina E, Ax A, Musumeci O, Longo M, Granata F. Posterior reversible encephalopathy syndrome (PRES) and infection: a systematic review of the literature. Neurol Sci 2019;40(5):915–22.

28. Franceschi AM, Ahmed O, Giliberto L, Castillo M. Hemorrhagic Posterior Reversible Encephalopathy Syndrome as a Manifestation of COVID-19 Infection. American Journal of Neuroradiology [Internet] 2020 [cited 2020 Jun 1];Available from: http://www.ajnr.org/content/early/2020/05/21/ajnr.A6595

29. Moore JB, June CH. Cytokine release syndrome in severe COVID-19. Science 2020;368(6490):473–4.

30. Hayashi M, Sahashi Y, Baba Y, Okura H, Shimohata T. COVID-19-associated mild encephalitis/encephalopathy with a reversible splenial lesion. J Neurol Sci 2020;415:116941.

31. Vellieux G, Rouvel-Tallec A, Jaquet P, Grinea A, Sonneville R, d’Ortho M-P. COVID-19 associated encephalopathy: Is there a specific EEG pattern? Clinical Neurophysiology 2020;131(8):1928–30.

32. Vespignani H, Colas D, Lavin BS, et al. Report of EEG Finding on Critically Ill Patients with COVID □19. Ann Neurol 2020;ana.25814.

33. Oxley TJ, Mocco J, Majidi S, et al. Large-Vessel Stroke as a Presenting Feature of Covid-19 in the Young. N Engl J Med 2020;e60.

34. Beyrouti R, Adams ME, Benjamin L, et al. Characteristics of ischaemic stroke associated with COVID-19. J Neurol Neurosurg Psychiatry 2020;jnnp-2020–323586.

35. Yaghi S, Ishida K, Torres J, et al. SARS2-CoV-2 and Stroke in a New York Healthcare System. Stroke 2020;STROKEAHA120030335.

36. Toscano G, Palmerini F, Ravaglia S, et al. Guillain–Barré Syndrome Associated with SARS-CoV-2. N Engl J Med 2020;NEJMc2009191.

37. Zhao H, Shen D, Zhou H, Liu J, Chen S. Guillain-Barré syndrome associated with SARS-CoV-2 infection: causality or coincidence? The Lancet Neurology 2020;19(5):383–4.

38. Padroni M, Mastrangelo V, Asioli GM, et al. Guillain-Barré syndrome following COVID-19: new infection, old complication? J Neurol [Internet] 2020 [cited 2020 May 6];Available from: http://link.springer.com/10.1007/s00415-020-09849-6

39. Pfefferkorn T, Dabitz R, von Wernitz-Keibel T, Aufenanger J, Nowak-Machen M, Janssen H. Acute polyradiculoneuritis with locked-in syndrome in a patient with Covid-19. J Neurol 2020;

40. Alberti P, Beretta S, Piatti M, et al. Guillain-Barré syndrome related to COVID-19 infection. Neurol Neuroimmunol Neuroinflamm 2020;7(4):e741.

41. Bigaut K, Mallaret M, Baloglu S, et al. Guillain-Barré syndrome related to SARS-CoV-2 infection. Neurol Neuroimmunol Neuroinflamm 2020;7(5):e785.

42. Gigli GL, Bax F, Marini A, et al. Guillain-Barré syndrome in the COVID-19 era: just an occasional cluster? J Neurol 2020;

43. Dinkin M, Gao V, Kahan J, et al. COVID-19 presenting with ophthalmoparesis from cranial nerve palsy. Neurology 2020;

44. Gutiérrez-Ortiz C, Méndez A, Rodrigo-Rey S, et al. Miller Fisher Syndrome and polyneuritis cranialis in COVID-19. Neurology 2020;10.1212/WNL.0000000000009619.

45. Lechien JR, Chiesa-Estomba CM, De Siati DR, et al. Olfactory and gustatory dysfunctions as a clinical presentation of mild-to-moderate forms of the coronavirus disease (COVID-19): a multicenter European study. Eur Arch Otorhinolaryngol 2020;

46. Decavel P, Petit C, Tatu L. Tapia syndrome at the time of the COVID-19 pandemic: Lower cranial neuropathy following prolonged intubation. Neurology 2020;

